# Safety and implementation of a phase 1 randomized GLA-SE-adjuvanted CH505TF gp120 HIV vaccine trial in newborns

**DOI:** 10.1101/2024.10.15.24315548

**Authors:** Avy Violari, Kennedy Otwombe, William Hahn, Shiyu Chen, Deirdre Josipovic, Vuyelwa Baba, Asimenia Angelidou, Kinga K. Smolen, Ofer Levy, Nonhlanhla N Mkhize, Amanda S Woodward, Troy M. Martin, Bart Haynes, Wilton B. Williams, Zachary K. Sagawa, James Kublin, Laura Polakowski, Margaret Brewinski Isaacs, Catherine Yen, Georgia Tomaras, Lawrence Corey, Holly Janes, Glenda Gray

## Abstract

**Background:** The neonatal immune system is uniquely poised to generate broadly neutralizing antibodies (bnAbs) and thus infants are ideal for evaluating HIV vaccine candidates. We present the design and safety of a novel glucopyranosyl lipid A (GLA)-stable emulsion (SE) adjuvant admixed with a first-in-infant CH505 transmitter-founder (CH505TF) gp120 immunogen designed to induce precursors for bnAbs against HIV.

**Methods:** HVTN 135 is a phase I randomized, placebo-controlled trial of CH505TF+GLA-SE or placebo. Healthy infants in South Africa aged ≤5 days, born to mothers living with HIV but HIV nucleic acid negative at birth were randomized to five doses of CH505TF + GLA-SE or placebo at birth and 8, 16, 32, and 54 weeks.

**Results:** 38 infants (median age = 4 days; interquartile range 4, 4.75 days) were enrolled November 2020 to January 2022. Among 28 (10) infants assigned to receive CH505TF + GLA-SE (placebo), most (32/38) completed the 5-dose immunization series and follow-up (35/38). Solicited local and systemic reactions were more frequent in vaccine (8, 28.6% local; 16, 57.1% systemic) vs. placebo recipients (1, 10% local, p = 0.25; 4, 40.0% systemic, p = 0.38). All events were Grade 1 except two Grade 2 events (pain, lethargy). Serious vaccine-related adverse events were not recorded.

**Conclusions:** This study illustrates the feasibility of conducting trials of novel adjuvanted HIV vaccines in HIV-exposed infants receiving standard infant vaccinations. The safety profile of the CH505TF + GLA-SE vaccine was reassuring.

**Trial registration:** ClinicalTrials.gov NCT04607408

**Funding:** National Institute of Allergy and Infectious Diseases (NIAID) at the National Institutes of Health (NIH)

**Brief summary:** This paper summarizes the phase 1 trial design and safety profile of an experimental CH505TF immunogen + GLA-SE HIV vaccine in infants born to mothers living with HIV.

## INTRODUCTION

Standard clinical development for vaccines with potential applications in infants generally involves early phase evaluations in adults, followed by evaluation in older children, and eventually neonatal populations, also called the “age de-escalation” approach; however, this process can take decades. In the case of preventive HIV vaccines, early evaluation in newborns and infants is supported by the following rationale: (a) a growing literature suggests that due to a distinct immunity profile in early life, natural HIV acquisition or immunization may more readily and rapidly induce broadly neutralizing antibodies (bnAbs) in infants than in adults (1–3); (b) infants may be an ideal population for current vaccine strategies that will likely require multiple doses administered at long intervals prior to achieving protective immune responses because, despite an initial period of risk of HIV-acquisition associated with breastfeeding, infants are likely to have a long period with low risk of HIV exposure prior to sexual debut; and (c) Infant populations already receive many vaccinations throughout the first year of life as part of standard of care, reducing the downstream operational complexities of implementing a potentially complex bnAb vaccine regimen.

The Antibody Mediated Prevention (AMP) trials (HVTN 703/HPTN 081 and HVTN 704/HPTN 085) provide proof that bnAbs can prevent HIV infection (4). Given this, we reason that neonatal populations should be integrated into the early clinical development plan of preventive HIV vaccines because vaccine candidates that might not elicit bnAbs in adults or older children could plausibly do so in infants (1–3). Furthermore, vaccine immunogenicity and safety in early life is not directly translatable from adult studies. While the neonatal immune system has been traditionally described as “immature”, or naïve, current understanding is that it is “distinct”, characterized by an initial fetal and maternal immune tolerant state with CD4 Th2 polarized responses and underdeveloped germinal centers (5), such that immune responses to pathogens and vaccines differ from those of adults. A broad range of relative immunogenicity is observed in newborns compared to older children and adults for various vaccines. For example, pneumococcal polysaccharide vaccine is not immunogenic in children less than two years of age (6), the DTP vaccine exhibits reduced immunogenicity (7), and the pneumococcal conjugate vaccine (PCV) has demonstrated comparable binding antibody responses with higher avidity (8, 9). Indeed, some measures of antibody quality, such as avidity, were higher in infants vaccinated with PCV7 at birth when compared to standard Expanded Program on Immunization (EPI) vaccine schedules beginning at six weeks of age (8).

HVTN 135 was designed to assess the HIV envelope CH505TF immunogen adjuvanted with GLA-SE in neonates. The immunogen was designed to induce antibody lineages for the CD4 binding site that have the potential to develop neutralization capacity. The addition of a novel adjuvant may be central to enhancing vaccine immunogenicity in vulnerable populations, including newborns and young infants. Many early studies use Alum, the most common vaccine adjuvant included in most licensed pediatric vaccines. Although Alum is generally safe, there are known limitations to the effectiveness of this Th2-polarizing adjuvant in infant populations (10). GLA-SE is a synthetic TLR4 agonist formulated in a nano-emulsion of squalene oil, is Th1 polarizing, and was shown to increase antibody avidity and breadth when compared to Alum in infant macaques (10). A key goal of this study was to establish the safety of this novel adjuvant in human infants.

Conducting early phase clinical trials in neonatal populations is logistically challenging due to the inherent vulnerability of this population. Infants exposed to HIV are at higher risk for poor outcomes relative to neonates who are not exposed (11), most often due to other concurrent infections, particularly lower respiratory tract infections, gastroenteritis, and sepsis. Establishing a framework to conduct early phase vaccine trials in a safe and ethical manner in this vulnerable population is therefore of critical importance.

To this end, a team of African- and US-based investigators, community representatives and other stakeholders convened to design and conduct a phase 1 clinical trial to test the safety, tolerability, and immunogenicity of a novel GLA-SE-adjuvanted CH505TF gp120 vaccine in healthy newborn infants exposed to HIV and living without HIV in South Africa. Here we report the safety data and provide insights on preparatory activities, study design elements and implementation considerations from this phase 1 trial, the first of its kind conducted in the last decade among infants exposed to HIV who did not acquire HIV.

## RESULTS

### Demographics and Baseline Characteristics

Among 63 mothers who provided consent for themselves and gave permission for their infants to participate in the study, 46 were assessed for eligibility within 5 days of birth (17 became ineligible during pregnancy), and 38 mother/infant pairs were enrolled (**Figure 1**). Most screening failures were due to maternal ineligibility (17 of 25 who were not enrolled), with the most common reasons being acute medical incidents, including COVID-19 (**Figure 1**).

**Figure 1.**
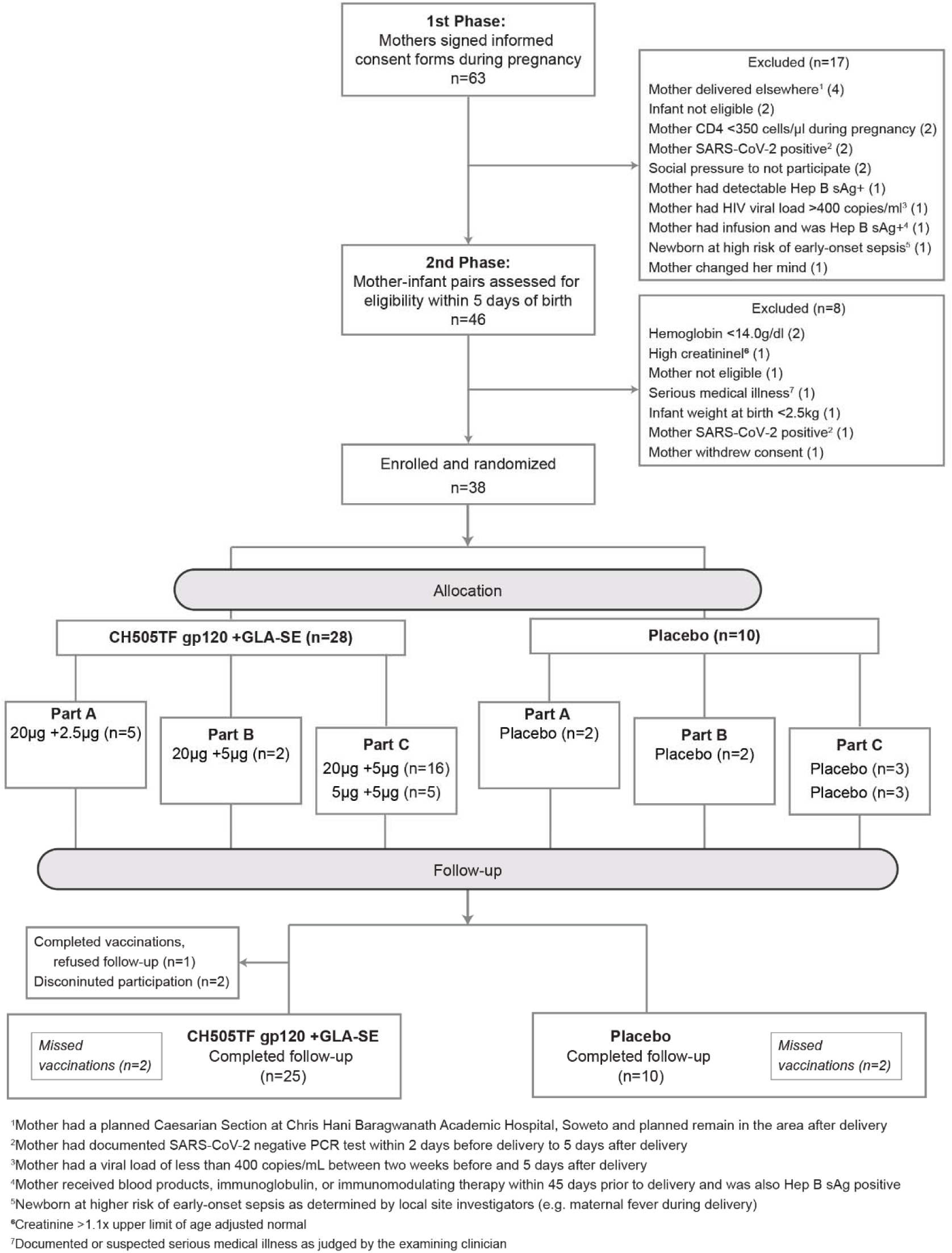
CONSORT diagram. Flow of participants, from screening to randomization, enrolment, follow-up, and study completion.

Of 38 infants enrolled, 18 (47.4%) were assigned female sex at birth (**Table 1**). Median age at first dose of study product was 4 days. The median (Interquartile Range) infant weight at enrollment was 3,035 (2,742.5, 3,377.5) grams and all were receiving ART prophylaxis. Most infants were breastfed at birth although this declined by 6 months (**Supplemental Table 1**). The majority of mothers (71%) were between 31 and 40 years of age; 45% had some secondary/high school education and 40% had completed secondary/high school. All mothers had a viral load below 400 copies/ml, while 95% had a viral load below the lower limit of detection for the assay used (**Supplemental Table 2**). Maternal and infant baseline characteristics were well balanced between vaccine and placebo groups.

**Table 1.**
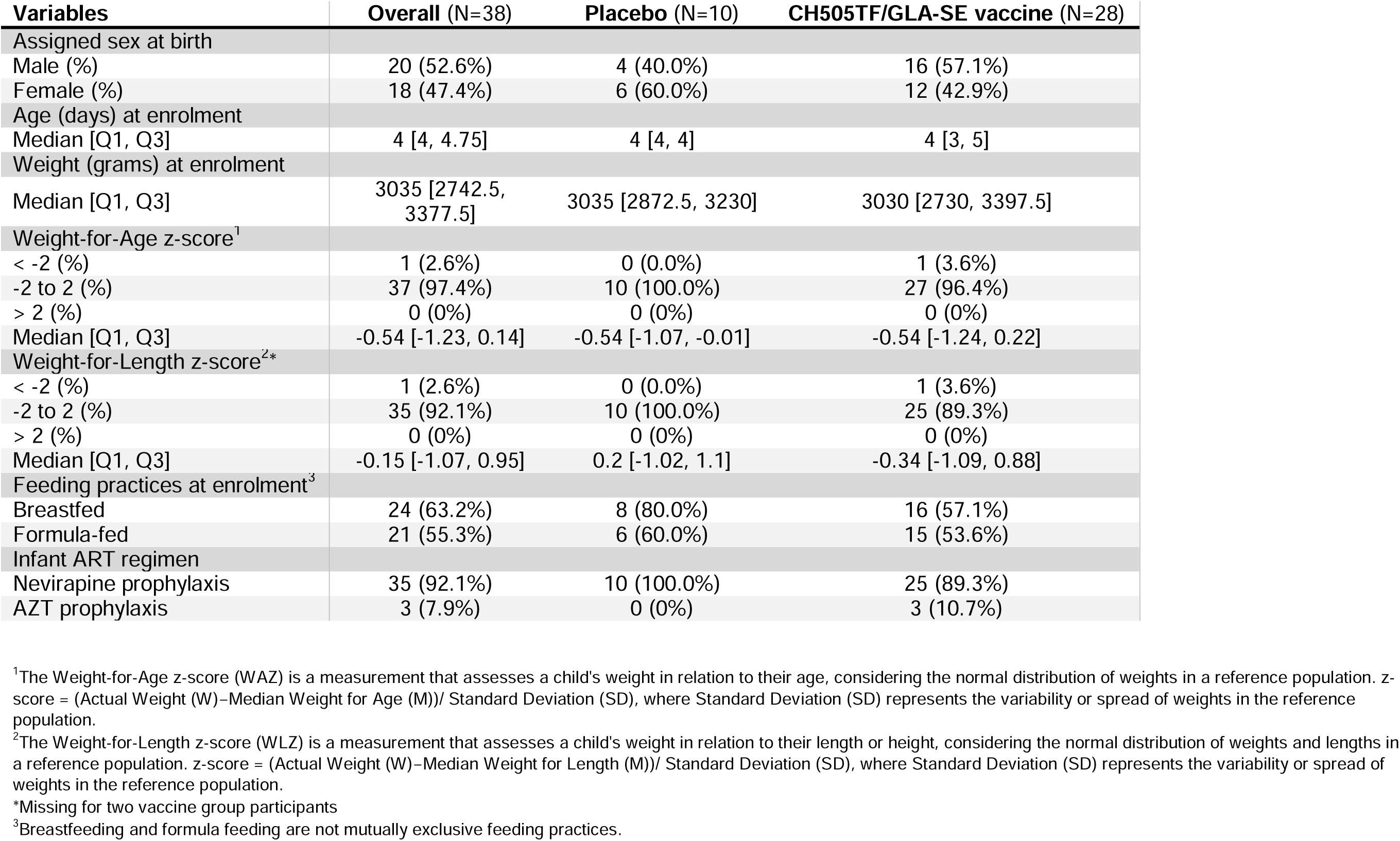
Infant participant characteristics at enrolment. Number and percent (or median and interquartile range) of infants enrolled, overall and by vaccine vs. placebo group.

| Variables | Overall (N=38) | Placebo (N=10) | CH505TF/GLA-SE vaccine (N=28) |
| --- | --- | --- | --- |
| Assigned sex at birth |  |  |  |
| Male (%) | 20 (52.6%) | 4 (40.0%) | 16 (57.1%) |
| Female (%) | 18 (47.4%) | 6 (60.0%) | 12 (42.9%) |
| Age (days) at enrolment |  |  |  |
| Median [Q1, Q3] | 4 [4, 4.75] | 4 [4, 4] | 4 [3, 5] |
| Weight (grams) at enrolment |  |  |  |
| Median [Q1, Q3] | 3035 [2742.5, 3377.5] | 3035 [2872.5, 3230] | 3030 [2730, 3397.5] |
| Weight-for-Age z-score<sup>1</sup> |  |  |  |
| < -2 (%) | 1 (2.6%) | 0 (0.0%) | 1 (3.6%) |
| -2 to 2 (%) | 37 (97.4%) | 10 (100.0%) | 27 (96.4%) |
| > 2 (%) | 0 (0%) | 0 (0%) | 0 (0%) |
| Median [Q1, Q3] | -0.54 [-1.23, 0.14] | -0.54 [-1.07, -0.01] | -0.54 [-1.24, 0.22] |
| Weight-for-Length z-score<sup>2*</sup> |  |  |  |
| < -2 (%) | 1 (2.6%) | 0 (0.0%) | 1 (3.6%) |
| -2 to 2 (%) | 35 (92.1%) | 10 (100.0%) | 25 (89.3%) |
| > 2 (%) | 0 (0%) | 0 (0%) | 0 (0%) |
| Median [Q1, Q3] | -0.15 [-1.07, 0.95] | 0.2 [-1.02, 1.1] | -0.34 [-1.09, 0.88] |
| Feeding practices at enrolment<sup>3</sup> |  |  |  |
| Breastfed | 24 (63.2%) | 8 (80.0%) | 16 (57.1%) |
| Formula-fed | 21 (55.3%) | 6 (60.0%) | 15 (53.6%) |
| Infant ART regimen |  |  |  |
| Nevirapine prophylaxis | 35 (92.1%) | 10 (100.0%) | 25 (89.3%) |
| AZT prophylaxis | 3 (7.9%) | 0 (0%) | 3 (10.7%) |

<sup>1</sup>The Weight-for-Age z-score (WAZ) is a measurement that assesses a child's weight in relation to their age, considering the normal distribution of weights in a reference population. z-score = (Actual Weight (W)–Median Weight for Age (M))/ Standard Deviation (SD), where Standard Deviation (SD) represents the variability or spread of weights in the reference population.
<sup>2</sup>The Weight-for-Length z-score (WLZ) is a measurement that assesses a child's weight in relation to their length or height, considering the normal distribution of weights and lengths in a reference population. z-score = (Actual Weight (W)–Median Weight for Length (M))/ Standard Deviation (SD), where Standard Deviation (SD) represents the variability or spread of weights in the reference population.
\*Missing for two vaccine group participants
<sup>3</sup>Breastfeeding and formula feeding are not mutually exclusive feeding practices.

### Compliance and Retention

Study retention was robust (**Supplemental Table 3**). Of the 38 infants enrolled, 35 completed follow-up. For one vaccine recipient, after completing vaccinations the caregiver refused follow-up. Another vaccine recipient was lost to follow-up when the caregiver relocated following the second vaccination, and the caregiver of third vaccine recipient refused further participation for the infant following the third vaccination. Receipt of the five-dose regimen was complete for 32 of 38 participants (24 of 28 vaccine recipients; 8 of 10 placebo recipients; **Figure 1**). Of the 6 participants who did not receive all five doses, two vaccine recipients mentioned above were due to early study termination while the rest were due to missed visits during the study None of the vaccine recipients missed vaccinations or terminated from the study early because of study product-related AEs.

### Safety

The vaccine demonstrated an acceptable safety profile and was well-tolerated (**Figure 3**, **Supplemental Table 4** and **Supplemental Figure 1**). Both local and systemic reactogenicity was typically mild. Among all participants, the most common solicited AEs were sleepiness/lethargy (23.7%) and rash (26.3%), while mild fever was seen in 15.8%. One moderate severity event of pain/tenderness occurred in Group 5 (CH505TF gp120 20μg/GLA-SE 5μg) after the third dose and one moderate sleepiness/lethargy event occurred in Group 7 (CH505TF gp120 5μg/GLA-SE 5μg) after the first dose. Neither required medical intervention. No severe or life-threatening systemic or local solicited AEs were observed. Local reactogenicity rates were higher in vaccine vs. placebo groups (p = 0.25, local), whereas the rates of systemic solicited AEs were similar between groups (p = 0.38).

Unsolicited AEs were generally mild or moderate in severity and consistent with common childhood illnesses (**Supplemental Table 5**). Six AEs that met seriousness criteria (SAEs) were reported: 3 in the placebo group and 3 in the vaccine group (**Table 2b**), all of which were deemed unrelated to the study product. Only 1 unsolicited AE (a grade one decreased neutrophil count) was deemed related to study product; this occurred in the placebo arm (**Supplemental Table 6**). Decreased neutrophil count not deemed related to study product was observed in all groups with approximately equivalent frequencies in each group and the highest numerical rate in the placebo arm (6/10 participants). None of the enrolled infants acquired HIV based on testing through a minimum of 711 days of life.

**Table 2a.** Most frequent adverse event summary. Adverse events occurring in 20% or more participants are shown. Number and percent of infants who experienced each adverse event, overall, by arm and grade of event.

| Event<br>N (%) | Overall<br>(N=38) | Placebo (N=10) |  |  | CH505TF/GLA-SE vaccine (N=28) |  |  |
| --- | --- | --- | --- | --- | --- | --- | --- |
|  |  | Grade 1 | Grade 2 | Grade 3 | Grade 1 | Grade 2 | Grade 3 |
| Lower respiratory tract infection | 4 (10.5%) | . | . | 1 (10.0%) | . | 2 (7.1%) | 1 (3.6%) |
| Upper respiratory tract infection | 10 (26.3%) | 1 (10.0%) | 1 (10.0%) | . | 3 (10.7%) | 5 (17.9%) | . |
| Gastrointestinal tract | 18 (47.4%) | 6 (60.0%) | 1 (10.0%) | . | 7 (25.0%) | 4 (14.3%) | . |
| Skin infections | 6 (15.8%) | 1 (10.0%) | . | 1 (10.0%) | 3 (10.7%) | 1 (3.6%) | . |
| Conjunctivitis | 3 (7.9%) | 1 (10.0%) | 1 (10.0%) | . | 1 (3.6%) | . | . |
| Underweight | 9 (23.7%) | 4 (40.0%) | . | . | 5 (17.9%) | . | . |
| Presumed allergic | 1 (2.6%) | . | . | . | 1 (3.6%) | . | . |
| Haematological abnormalities | 30 (78.9%) | 4 (40.0%) | 5 (50.0%) | . | 17 (60.7%) | 4 (14.3%) | . |
| CMV Hepatitis | 1 (2.6%) | 1 (10.0%) | . | . | . | . | . |
| Sepsis | 1 (2.6%) | . | . | . | . | . | 1 (3.6%) |

**LRTI:** includes bronchiolitis, bronchitis, pneumonia; **URTI:** includes pharyngitis, nasopharyngitis, tonsillitis, otitis;
**Gastrointestinal:** includes gastroenteritis, diarrhoea, vomiting; **Skin infections:** tinea corporis, impetigo, fungal infection

**Table 2b.** Serious adverse event summary. Number and percent of participants who experienced serious adverse events (SAEs), overall, by arm and grade of event. All events were determined to be unrelated to study product.

| Event | Overall<br>(N=38) | Placebo (N=10) |  | CH505TF/GLA-SE vaccine<br>(N=28) |  | Days since last<br>injection |
| --- | --- | --- | --- | --- | --- | --- |
| N (%) |  | Grade 3 | Grade 4 | Grade 3 | Grade 4 |  |
| Blood and lymphatic<br>system disorders |  |  |  |  |  |  |
| Lymphadenitis | 1 (2.6%) | 1 (10.0%) | . | . | . | 21 |
| Infections and<br>infestations |  |  |  |  |  |  |
| Bronchiolitis | 1 (2.6%) | . | . | 1 (3.6%) | . | 50 |
| Fungal skin infection | 1 (2.6%) | 1 (10.0%) | . | . | . | 43 |
| Sepsis | 1 (2.6%) | . | . | 1 (3.6%) | . | 12 |
| Ophthalmia neonatorum | 1 (2.6%) | . | . | 1 (3.6%) | . | 2 |
| Pneumonia | 1 (2.6%) | 1 (10.0%) | . | . | . | 72 |

### Early Infant Immunizations

BCG, RV, DTaP-IPV-Hib-HBV, and PCV vaccination rates were high (> 97%) through the first 9 months of life (**Supplemental Table 3**). OPV vaccine receipt was lower in frequency at birth (79%) and 6 weeks (84%). Ninety-seven percent of infants received measles vaccination at 12 months and 91% received DTaP-IPV-Hib-HBV at 18 months. The most common reason for missed vaccinations was out-of-stock product (88% of the 69.5% of missed vaccinations for which a reason was provided).

### Infant growth

At enrolment, 37 of 38 infants (97.4%) and 35 of 38 (92.1%) had weight-for-age and weight-for-length z-scores, respectively, within 2 standard deviations of the population mean (z-scores between −2 and 2). Healthy growth was observed in all infants over the two years of follow-up, with weight-for-age consistently within 2 standard deviations of the mean, and average weight-for-age trajectories following the population mean (**Supplemental Figure 2**). Vaccine and placebo weight-for-age and weight-for length averages were similar.

## DISCUSSION

Conducting vaccine trials in a neonatal population presents substantial scientific, ethical, and operational challenges. HVTN 135 provides reassuring safety data for the studied HIV vaccine that includes a novel adjuvant (GLA-SE) and demonstrates the feasibility and safe implementation of a Phase I study in neonates vaccinated within 5 days of birth. Additionally, the study demonstrates the feasibility of collecting cord blood samples that can be used for planned cellular analysis in the context of an HIV vaccine trial.

The safety and tolerability of the candidate vaccine were reassuring. There were only two moderate (Grade 2) vaccine reactogenicity events, one for pain/tenderness at the injection site in the 20ug CH505TF gp120 5ug GLA-SE group and another for sleepiness/lethargy in the 5ug CH505TF gp120 5ug GLA-SE group. All other reactogenicity events were mild. Overall, 61% of participants experienced a reactogenicity event. All unsolicited adverse events were deemed not related to study product, except for one Grade 1 neutropenia in the placebo group which had been assessed as related prior to unblinding. Neutropenia was common in our study with an overall rate of 34% (60% in the placebo group and 25% in the vaccine group) and was primarily attributed to Cotrimoxazole prophylaxis which was part of the standard of care for infants exposed to HIV at the time of enrollment. Mild neutropenia is a commonly observed laboratory abnormality in this population, with high rates of neutropenia occurring in previous vaccine trials in infants exposed to HIV mostly attributed to concomitant medications such as Cotrimoxazole (18).

Neonatal and infant vaccination is one of the most important interventions in child health - often delivered at large scale and at the primary health care level. This study opens the possibility for similar vaccine trials against HIV and other pathogens in this population. Further studies of candidate HIV vaccines with similarly acceptable safety and feasibility would provide support for including infants in trials at a much earlier stage in vaccine development and may accelerate vaccine development and enhance vaccine availability for younger populations.

Choosing infants exposed to HIV as the study population has several advantages and disadvantages. The same population has been involved in previous HIV vaccine studies and it is conceivable that acceptability of such a study may be higher in families affected by HIV. However, interpretation of immune responses to the vaccine can be more challenging by the presence of maternal antibodies (anti gp 120 and anti-CD4bs) which are present for at least 6 months. Co-enrolling the mother in the study and including a placebo arm facilitates interpretation of the results.

Any Phase I study in this vulnerable age group should consider input from experts in neonatal immunology and vaccinology as well as include consultation with ethicists and community advisory members. Our study was successful in incorporating community feedback throughout protocol development; importantly, this began very early during the protocol design phase. By engaging with the community early in the process, we built trust and maintained open channels of communication, which resulted in robust recruitment and retention. For example, in addition to the care related to vaccination, the community team highlighted that an open-door policy with access to routine pediatric care as needed would also be an important element for participation in the study for women and their infants. The community team also engaged community members on the acceptability of cord blood donation. We would recommend that other early-phase vaccine studies utilize similar methods to ensure community involvement.

Limiting inclusion to Caesarian deliveries allows for collection of cord blood allowing for in depth immunologic testing and sparing the amount of blood taken from the baby at birth. It could however slow down enrollment either by limiting the pool of mothers available for recruitment or when participants deliver vaginally prior to the scheduled Caesarian delivery, as it was the case in our study. Planned C/S in low-resource areas can often take place outside the scheduled date due to multiple obstetric emergencies that take precedent. This requires the study team and the laboratory receiving the specimens to be on standby and available at night, which requires resources and logistics that in the case of our study were further complicated by the COVID pandemic and subsequent frequent restrictions at operating theatres.

An important consideration for our study is that healthy, neonates exposed to HIV and living without HIV have been shown to have a higher rate of viral infections and have a higher burden of serious health outcomes than their HIV-unexposed counterparts (19, 20). Including HIV-exposed infants in a clinical trial with a rigorous structure for ensuring participant safety entailed careful monitoring of infant growth rate as well as key health outcomes. Reassuringly, we found that the infants in our study developed at a rate within the normal range. By including details of the protocol conduct, we provide the wider pediatric community insight into the operational details of a neonatal vaccine trial in a population of infants exposed to HIV. Another benefit of our trial is that robust, high quality safety data from the placebo arm can help to inform the design of future studies in this population.

During protocol development it was also critical to factor in that infants have substantial limitations on sampling that can be performed, especially with respect to blood volumes. Substantial limitations on blood volumes require careful planning for the immunologic specimens that can be collected, and careful monitoring for iatrogenic anemia. To make the best use of the small sample volume available, experts in immunology and vaccinology were engaged to design an experimental plan that leveraged novel platforms and cutting-edge technology. Ongoing work will examine molecular signatures underlying the observed immunogenicity effects using modern systems vaccinology approaches. Evaluation of the immunogenicity of the current vaccine is also ongoing, including a planned comparison of infants in HVTN 135 with adults in HVTN 115 receiving the same GLA-SE adjuvanted gp120 protein vaccine, which will offer unique insight into the adult versus infant immune response to this vaccine. Despite the challenges of conducting research in a neonatal population, there is an urgent need to study the molecular mechanisms of vaccine safety and immunogenicity in this age group, which has distinct immunologic features and receives many vaccines.

In addition to the expected scientific insights, HVTN 135 serves as an example for integrating neonatal vaccines early into the vaccine development plan and its operational conduct provides a template for future vaccine trials conducted in this population, including additional pediatric HIV vaccine trials. A successful neonatal vaccine could potentially be given to all infants irrespective of whether their birth parent is living with HIV, supporting a continuum of protection from early life to sexual debut.

## METHODS

### Ethical Considerations

Conducting vaccine research in infants presents several challenging ethical issues related to parental permission, risks and benefits, and blood draw limits. The HIV Vaccine Trials Network (HVTN) incorporated teams with expertise in neonatal immunity, and systems vaccinology. The protocol included sample sparing assays, and the team conducted extensive stakeholder engagement with community representatives and with ethical and regulatory consultants during protocol development. The trial was overseen by the University of the Witwatersrand Human Research Ethics Committee (HREC), reference number 190914B.

### HVTN 135 Study Design

#### Study Population

Participants were healthy newborn infants exposed to HIV who were born to mothers living with HIV in South Africa, enrolled within 5 days of birth. Key inclusion criteria for the pregnant participants were well-controlled HIV, as demonstrated by virologic suppression (HIV viral load of less than 400 copies/mL) and a CD4 count >350 cells/microliter, and who planned to have Caesarean delivery. Planned Caesarean delivery was required due to the need to collect cord blood which is logistically challenging in the setting of spontaneous vaginal delivery. Key inclusion criteria for the infant included a birth weight above 2.5kg, estimated gestational age >37 weeks, a negative HIV nucleic acid test at birth, and antiretroviral prophylaxis consistent with the local standard of care.

#### Sex as a biological variable

Our study examined male and female infants and no differences in safety profile were detected between the sexes.

#### Study Investigational Products

The CH505TF gp120 immunogen was derived from a clade C transmitted/founder HIV virus isolated from a single acutely HIV–infected donor from Malawi (CH505) (12–14). The adjuvant GLA-SE is an oil-in-water stable emulsion containing the immunological adjuvant Glucopyranosyl Lipid A (GLA). CH505TF gp120 is manufactured by Berkshire Sterile Manufacturing and GLA-SE is manufactured by the Infectious Disease Research Institute (IDRI); both products were provided by DAIDS.

#### Design and Rationale

A three-part randomized, placebo-controlled design was used (**Figure 1** and **Figure 2**). A placebo group was deemed important to aid unbiased reporting and interpretation of safety events. Given that infants who are exposed to HIV acquire HIV-specific immune responses from their mothers, including a placebo arm also allowed differentiating vaccine-induced versus maternally-derived immune responses. The three-part design minimized exposure to higher doses of the adjuvant until safety data with lower doses were collected. Part A of the study provided initial safety data on the CH505TF immunogen (20μg) when combined with a 2.5μg dose of the GLA-SE adjuvant. Part B provided a preliminary safety assessment of the maximum targeted 5 mcg dose of the GLA-SE adjuvant, since this dose was under concurrent evaluation in separate non-HIV-vaccine pediatric studies (NCT03806699 and NCT03799510). Enrollment into Part B began after the study’s first planned safety hold and review of the cumulative safety data for all participants in Part A up to and including two weeks following the first dose of study product (i.e., CH505TF+GLA-SE or placebo). Part C expanded the evaluation of the 20μg CH505TF/ 5μg GLA-SE regimen, and also included for comparison a dose-sparing 5μg CH505TF + 5μg GLA-SE arm. Enrollment into Part C began after a second planned safety hold and review of cumulative safety data for all participants in Part A and for all participants in Part B up to and including two weeks following the third dose of study product.

**Figure 2.**
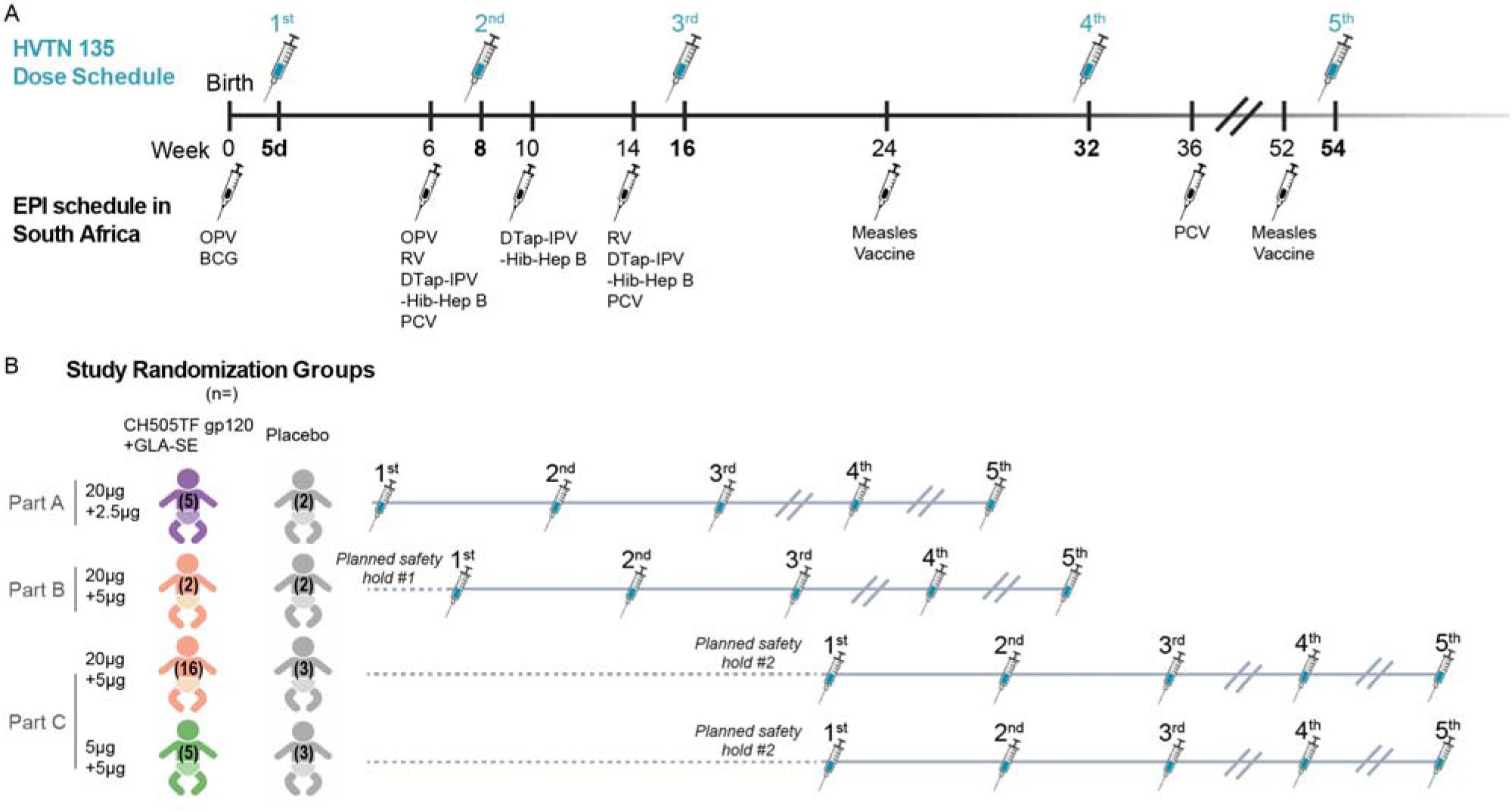
Study design. (A) Vaccine dosing schedule in relation to EPI vaccinations. (B) Study Schema: Healthy infants without HIV born to mothers living with HIV were randomized to receive 5 doses of CH505TF gp120 + GLA-SE vaccine or placebo, with the first dose within 5 days of birth. The 3-part design allowed for an initial assessment of safety in Part A before the target adjuvant dose was studied in Part B. Part C compared two different doses of CH505TF gp120 while keeping the same adjuvant dose.

In each part, participants were randomized to receive five doses of the vaccine or placebo, at Weeks 0, 8, 16, 32, and 54. This schedule was designed to minimize overlap with the Republic of South Africa (RSA) EPI vaccination schedule (see **Figure 2A**). If an infant was found to be delayed in EPI vaccines at a study visit, priority was given to catching up on the EPI vaccines before administering study product. EPI vaccines were given at least 7 days prior to administration of study vaccination.

**Figure 3.**
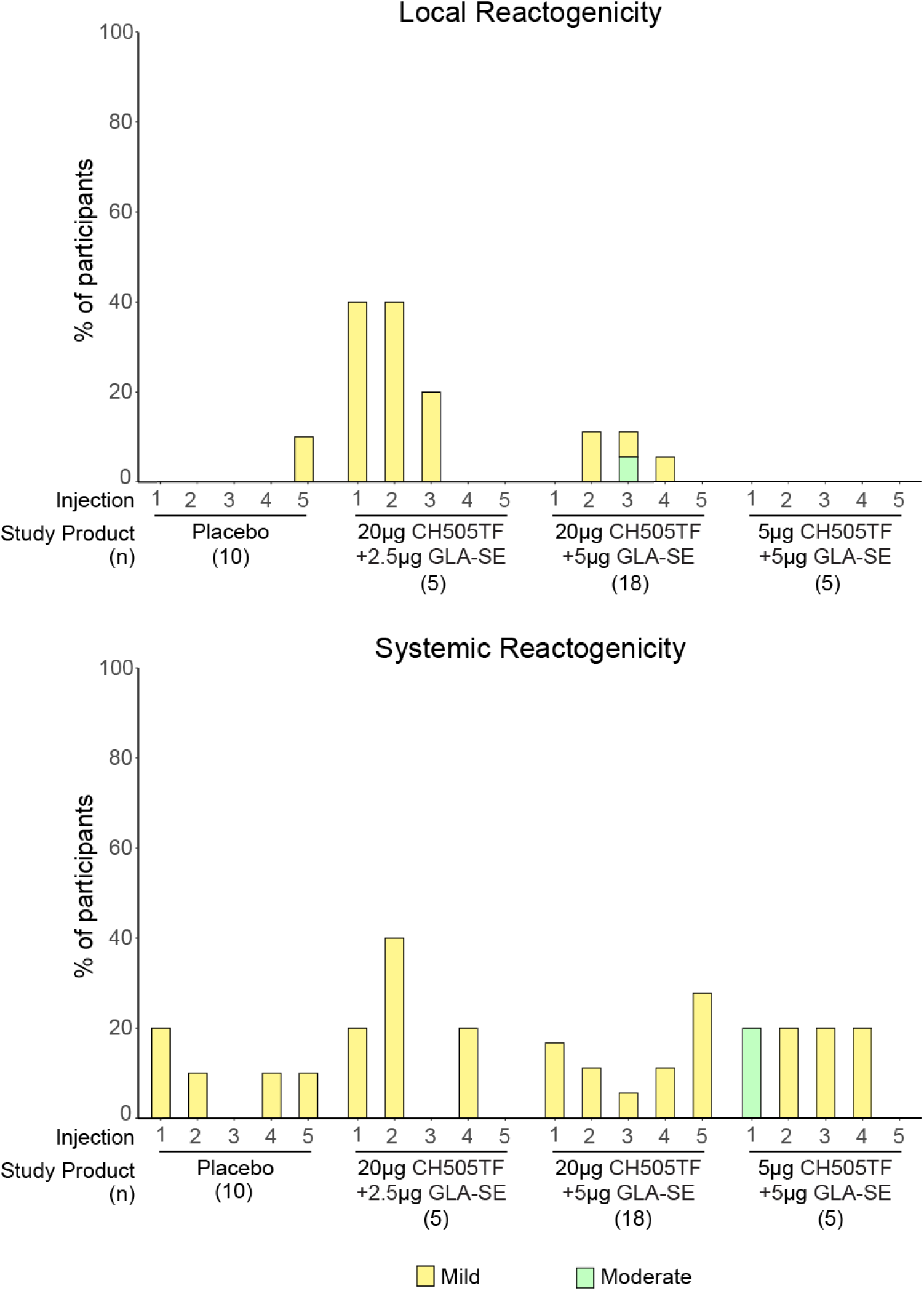
Local and systemic reactogenicity. Percentage of participants who experienced each grade of local and systemic reactogenicity by arm.

#### Study Procedures

Participants received study product at the indicated doses via 0.25mL or 0.5mL injections administered intramuscularly in the thigh. Both the placebo and the diluent were 0.9% Sodium Chloride. All participants were observed for a minimum of 60 minutes following study product administration.

For seven days following each study product injection, caregivers recorded pre-defined solicited adverse events (AEs) (also known as “reactogenicity”) using a participant diary. Unsolicited AEs were coded per the Medical Dictionary for Regulatory Activities (MedDRA) and graded for severity according to the Division of AIDS (DAIDS) Table for Grading of the Severity of Adult and Pediatric Adverse Events, version 2.1. Participants were followed for 12 months following the final study product dose, through approximately two years of life, with a total of 14 in-person visits (see **Study Protocol**).

Protocol-required samples included cord blood and peripheral infant blood specimens for safety and immunogenicity analyses. Optional samples included maternal breast milk, and stool from both mother and infant. Infant anthropometric measurements were obtained at each study visit to monitor infant growth as part of ongoing safety assessments and in relation to World Health Organization (WHO) growth standards (15).

Infants were PCR-tested for HIV at birth, 10 weeks, and 6 months per the RSA standard of care (SoC) (16). PCR tests were performed at well-baby clinics, with results provided to the clinical research site. The SoC requires anti-HIV antibody testing at 18 months of age for all infants. To distinguish between vaccine-induced antibodies and HIV acquisition, infants had blood collected for testing via an HVTN-developed diagnostic algorithm at Study Month 17, with results provided to the local pediatrician. Infants were also tested via the HVTN diagnostic algorithm at the final visit at Study Month 24.5 to rule out HIV acquisition during the course of follow-up.

Sample collections from the infant participants were designed to maximize the scientific information obtained from small volumes while ensuring the overall safety of the participants (17). This approach was complemented by routine monitoring of hemoglobin to monitor for anemia during the study.

### HVTN 135 Trial: Implementation

#### Site outreach to potential participants

Potential participants were recruited by the Perinatal HIV Research Unit (PHRU) at Chris Hani Baragwanath Hospital in Johannesburg, South Africa. Two factors informed the recruitment strategy. First was the requirement for delivery by Caesarean delivery (see Study Population section above), and second was to identify potential birth parents who would be able to follow the study schedule.

Since Caesarean deliveries are not performed as standard of care for pregnant individuals living with HIV in RSA, in collaboration with the Chris Hani Baragwanath Academic Hospital (CHBAH) Obstetrics Department, individuals likely to undergo Caesarian delivery (for example, with a previous Caesarean delivery) were approached for participation in the study.

Recruitment was conducted by professional nurses who had the requisite knowledge and understanding to explain and answer questions about the study and any complex concepts. Possible barriers to retention were addressed by ascertaining the pregnant individual’s current and future emotional and financial support network. With the mother’s consent, the nurses invited and engaged other individuals forming part of the mother’s support system and potentially involved with decisions regarding the future of the infant.

#### Maternal support

The mothers had open access to the PHRU clinic during weekdays, with the ability to come to the clinic any time they needed. At enrollment they were provided with a 24-hour emergency number for one of the study doctors. During the hospital admission for delivery, the study team had daily contact with the mothers and the CHBAH Obstetrics team. Two on-call study team members at minimum were present at the delivery, often consisting of two nurses, or sometimes one nurse and one doctor. After the birth, the team had daily contact with the mother and the CHBAH staff caring for the infant.

A driver transported the mother and infant to and from the PHRU clinic for study visits during the first 2 to 4 weeks post-partum and whenever needed thereafter. On site counselling services were made available for the mothers, which offered psychological support. Nutritional support and iron supplementation were provided as needed for the infants.

### Navigating the COVID-19 pandemic

The study enrolled participants during the COVID-19 pandemic between November 2020 and July 2022, representing an additional challenge to this complex study. The study team adhered to the PHRU and CHBAH COVID-19 policies designed to minimize risks to staff and participants. The protocol was amended to exclude birthing parents who tested positive for SARS-CoV-2 by PCR.

During periods of COVID-19 outbreaks in the country, changes in the CHBAH Obstetrics Department to mitigate risk and manage patient burden and staff illnesses included allocating dedicated COVID-19 wards and operating theatres and referring patients without COVID-19 to other hospitals for Caesarean delivery. This meant that the study team on call had to be available 24 hours per day and had to follow up with participants in other hospitals, ensure equipment and laboratory kits were readily accessible, and organize 24-hour laboratory sample collection and processing.

### Statistical analysis

Frequencies and proportions were calculated for categorical variables, whereas medians and interquartile ranges were determined for continuous measures. WHO growth standards (15) were used to interpret weight-for-age and weight-for-length z-scores that were categorized into three groups: <-2 (indicative of underweight for weight-for age; wasting for weight-for length), −2 to 2 and > 2. Participants were analyzed by randomization group, except that all placebo recipients and the two cohorts from Part B and C that received the same vaccine dose were combined for analysis. Statistical analysis was conducted using the open-source R program version 4.0.4 (https://www.R-project.org).

## Role of the funding source

The funding source was the Division of AIDS (DAIDS) of the National Institute of Allergy and Infectious Diseases (NIAID) of the US National Institutes of Health (NIH). Members of the DAIDS helped to draft the initial manuscript, but analysis was conducted independently. The funder had no role in data collection or statistical analyses.

## Data sharing and availability

Participant follow-up is still underway. At study closeout, deidentified quality assured study data will be deposited at https://clinicaltrials.gov under accession number NCT04607408.

## Author contributions

AV, OL, KKS, WH, GT, TM, BH, NNM, JK, WW, ZS, MBI, KO, LC, HJ, GG assisted with primary HVTN135 study design. HJ and SC accessed and verified the underlying data. OL, AA, AWD, KKS, TM, HJ, CY read the draft manuscript for detailed editorial feedback. LC provided funding, study design, writing-review and editing AV, WH, HJ, KO, DS, SC drafted the manuscript.

## Acknowledgements

The team acknowledges the stellar work of the staff at the PHRU Clinical Research Site. The Precision Vaccines Program (PVP) at Boston Children’s Hospital (BCH) thanks the leadership of BCH including Drs. Wendy Chung, Nancy Andrews and Kevin Churchwell for their support. Without the participation of the infants and their caregivers this study would not have been possible. Figure 2 was created using BioRender.com.

## Funding

The trial was funded through National Institute of Allergy and Infectious Disease of the National Institutes of Health under Grants UM1 AI068614 [HVTN Leadership and Operations Center], UM1 AI068635 [HVTN Statistical and Data Management Center], and UM1 AI068618 [HVTN Laboratory Center]. The members of the Precision Vaccines Program are supported in part via U.S. NIH/NIAID contracts and grants including Adjuvant Development Program Contract HHSN272201800047C (OL and KKS), U19 AI168643 Immune Development in Early Life (IDEAL; OL, KKS & AA), mentored clinical scientist development award 1K08AI168487 (AA), as well as a grant from HVTN (OL and KKS).

## Declaration of interests

OL is a named inventor on patents relating to vaccine adjuvants and to human in vitro systems that model the safety and immunogenicity of adjuvants, vaccines and immunomodulators. He serves as a consultant to GSK and Hillevax and is a co-founder of Ovax, Inc. All other authors declare no conflict of interest.

## Notes

### Clinical Trial

NCT04607408

### Author Declarations

The trial was overseen by the University of the Witwatersrand Human Research Ethics Committee (HREC), reference number 190914B.

