## Supplemental Materials for "Safety and implementation of a phase 1 randomized GLA-SE-adjuvanted CH505TF gp120 HIV vaccine trial in newborns"

Supplemental Table 1. Infant feeding practices.

Supplemental Table 2. Maternal participant characteristics at enrolment.

Supplemental Table 3. Summary of participant compliance and study vaccination

Supplemental Table 4. Summary of local and systemic reactogenicity events.

Supplemental Table 5. Summary of adverse events, regardless of relationship to study product.

Supplemental Table 6. Summary of adverse events deemed related to study product(s)

Supplemental Figure 1. Individual reactogenicity plots

Supplemental Figure 2. Anthropometric measures during follow-up

**Supplemental Table 1. Infant feeding practices.** Number and percent of enrolled infants overall with each characteristic.

|  | **Breastfed^*^, no. (%)** | **Formula-fed^+^, no. (%)** | **Fed complementary foods^+-^, no. (%)** | **Permanent cessation of breastfeeding^&^, no. (%)** | **Clinical concern with breastfeeding^$^, no. (%)** |
| --- | --- | --- | --- | --- | --- |
| Enrollment (N=38) | 24 (63.2%) | 21 (55.3%) | - | - | 1 (2.6%) |
| Month 4.5 (N=35) | 14 (40.0%) | 20 (57.1%) | 6 (17.1%) | 6 (17.1%) | 0 (0.0%) |
| Month 13 (N=33) | 11 (33.3%) | 22 (66.7%) | 33 (100.0%) | 11 (33.3%) | 2 (6.1%) |

Number in each row is the number of enrolled participants who have not discontinued the study by the time of the visit and who attended the visit. Breastfeeding, formula feeding, and complementary foods are not mutually exclusive feeding practices.

*Fed breast milk by the biological mother since the last visit (2 weeks prior)

+ Fed formula since the last visit (2 weeks prior)

+- Fed any complementary foods since the last visit (2 weeks prior)

& Permanent cessation of breastfeeding at any time point up to and including the visit day

$ Clinical concern with feeding since the last visit (2 weeks prior)

**Supplemental Table 2. Maternal participant characteristics at enrolment.** Number and percent of enrolled mothers with each characteristic, overall and by arm.

| **Variables** | **Overall** (N=38) | **Placebo** (N=10) | **CH505TF/GLA-SE vaccine** (N=28) |
| --- | --- | --- | --- |
| Age (years) |  |  |  |
| 21-25 | 3 (7.9%) | 0 (0.0%) | 3 (10.7%) |
| 26-30 | 7 (18.4%) | 3 (30.0%) | 4 (14.3%) |
| 31-35 | 15 (39.5%) | 4 (40.0%) | 11 (39.3%) |
| 36-40 | 12 (31.6%) | 3 (30.0%) | 9 (32.1%) |
| 41-50 | 1 (2.6%) | 0 (0.0%) | 1 (3.6%) |
| Race |  |  |  |
| Black | 38 (100.0%) | 10 (100.0%) | 28 (100.0%) |
| Highest level of education |  |  |  |
| Completed graduate school (doctorate/masters/honours/higher education degree) | 1 (2.6%) | 0 (0.0%) | 1 (3.6%) |
| Completed secondary/high school | 15 (39.5%) | 2 (20.0%) | 13 (46.4%) |
| National certificate/trade certificate/national diploma/occupational certificate | 3 (7.9%) | 1 (10.0%) | 2 (7.1%) |
| Some secondary/high school | 17 (44.7%) | 6 (60.0%) | 11 (39.3%) |
| Some university/technical education (associates/bachelors/technical degree) | 2 (5.3%) | 1 (10.0%) | 1 (3.6%) |
| HIV viral suppression |  |  |  |
| Suppressed (detected but ≤400 copies/mL) | 2 (5.3%) | 1 (10.0%) | 1 (3.6%) |
| Undetectable (viral load not detected by test used) | 36 (94.7%) | 9 (90.0%) | 27 (96.4%) |
| Currently Have Medical Insurance |  |  |  |
| No | 38 (100.0%) | 10 (100.0%) | 28 (100.0%) |
| ART regimen |  |  |  |
| Efavirenz based regimen | 20 (52.6%) | 7 (70.0%) | 13 (46.4%) |
| Dolutegravir based regimen | 21 (55.3%) | 6 (60.0%) | 15 (53.6%) |
| Protease inhibitor | 2 (5.3%) | 0 (0.0%) | 2 (7.1%) |
| Rilpivirine based regimen | 1 (2.6%) | 0 (0.0%) | 1 (3.6%) |

Educational level and medical aid status is self-reported. HIV viral suppression as measured by qRT-PCR. Mothers may be on multiple ART regimens and therefore ART regimen percentages may be higher than 100%.

**Supplemental Table 3. Summary of participant compliance, study vaccination, and EPI vaccination.** Number and percent of enrolled participants with each characteristic, overall and by arm.

| **Variables** | **Overall**  **(n=38)** | **Placebo**  **(n=10)** | **20ug CH505TF gp120**  **2.5ug GLA-SE**  **(n=5)** | **20ug CH505TF gp120**  **5ug GLA-SE**  **(n=18)** | **5ug CH505TF gp120**  **5ug GLA-SE**  **(n=5)** |
| --- | --- | --- | --- | --- | --- |
| Participant status |  |  |  |  |  |
| Completed follow-up | 35 (92.1%) | 10 (100.0%) | 4 (80.0%) | 16 (88.9%) | 5 (100%) |
| Completed vaccinations, refused follow-up | 1 (2.6%) | 0 (0.0%) | 1 (20.0%) | 0 (0.0%) | 0 (0.0%) |
| Discontinued |  |  |  |  |  |
| Lost to follow-up | 1 (2.6%) | 0 (0.0%) | 0 (0.0%) | 1 (5.6%) | 0 (0.0%) |
| Withdrew consent | 1 (2.6%) | 0 (0.0%) | 0 (0.0%) | 1 (5.6%) | 0 (0.0%) |
| Number of vaccinations administered |  |  |  |  |  |
| Dose 1 (Week 0) | 38 (100.0%) | 10 (100.0%) | 5 (100.0%) | 18 (100.0%) | 5 (100.0%) |
| Dose 2 (Week 8) | 36 (94.7%) | 9 (90.0%) | 5 (100.0%) | 17 (94.4%) | 5 (100.0%) |
| Dose 3 (Week 16) | 35 (92.1%) | 9 (90.0%) | 5 (100.0%) | 17 (94.4%) | 4 (80.0%) |
| Dose 4 (Week 32) | 36 (94.7%) | 10 (100.0%) | 5 (100.0%) | 16 (88.9%) | 5 (100.0%) |
| Dose 5 (Week 54) | 35 (92.1%) | 10 (100.0%) | 5 (100.0%) | 15 (83.3%) | 5 (100.0%) |
| EPI vaccines administered |  |  |  |  |  |
| BCG at birth | 37/38 (97.4%) | 10/10 (100.0%) | 5/5 (100.0%) | 17/18 (94.4%) | 5/5 (100.0%) |
| OPV at birth | 30/38 (78.9%) | 7/10 (70.0%) | 5/5 (100.0%) | 15/18 (83.3%) | 3/5 (60.0%) |
| OPV at 6 weeks | 32/38 (84.2%) | 9/10 (90.0%) | 5/5 (100.0%) | 15/18 (83.3%) | 3/5 (60.0%) |
| RV at 6 weeks | 38/38 (100.0%) | 10/10 (100.0%) | 5/5 (100.0%) | 18/18 (100.0%) | 5/5 (100.0%) |
| DTaP-IPV-Hib-HBV at 6 weeks | 38/38 (100.0%) | 10/10 (100.0%) | 5/5 (100.0%) | 18/18 (100.0%) | 5/5 (100.0%) |
| PCV at 6 weeks | 38/38 (100.0%) | 10/10 (100.0%) | 5/5 (100.0%) | 18/18 (100.0%) | 5/5 (100.0%) |
| DTaP-IPV-Hib-HBV at 10 weeks | 36/37 (97.3%) | 9/10 (90.0%) | 5/5 (100.0%) | 17/17 (100.0%) | 5/5 (100.0%) |
| RV at 14 weeks | 36/37 (97.3%) | 10/10 (100.0%) | 5/5 (100.0%) | 16/17 (94.1%) | 5/5 (100.0%) |
| DTaP-IPV-Hib-HBV at 14 weeks | 36/37 (97.3%) | 10/10 (100.0%) | 5/5 (100.0%) | 16/17 (94.1%) | 5/5 (100.0%) |
| PCV at 14 weeks | 36/37 (97.3%) | 10/10 (100.0%) | 5/5 (100.0%) | 16/17 (94.1%) | 5/5 (100.0%) |
| Measles Vaccine at 6 months | 36/36 (100.0%) | 10/10 (100.0%) | 5/5 (100.0%) | 16/16 (100.0%) | 5/5 (100.0%) |
| PCV at 9 months | 36/36 (100.0%) | 10/10 (100.0%) | 5/5 (100.0%) | 16/16 (100.0%) | 5/5 (100.0%) |
| Measles Vaccine at 12 months | 35/36 (97.2%) | 10/10 (100.0%) | 4/5 (80.0%) | 16/16 (100.0%) | 5/5 (100.0%) |
| DTaP-IPV-Hib-HBV at 18 months | 32/35 (91.4%) | 8/10 (80.0%) | 4/4 (100.0%) | 15/16 (93.8%) | 5/5 (100.0%) |

The denominator for study completion, vaccination completion, and discontinuation is the number of enrolled infants. The denominator for EPI vaccine receipt is the number of infants under follow-up at the time of the scheduled vaccine. Loss to follow-up indicates that the site was unable to contact the mother or caregiver.

**Supplemental Table 4. Summary of local and systemic reactogenicity events.** Number and percent of enrolled participants who experienced each type of reactogenicity, overall and by arm, and by symptom severity.

| **Symptom**  **N (%)** | **Overall (N=38)** | **Placebo (N=10)** | **20ug CH505TF gp120**  **2.5ug GLA-SE**  **(N=5)** | **20ug CH505TF gp120**  **5ug GLA-SE**  **(N=18)** | **5ug CH505TF gp120**  **5ug GLA-SE**  **(N=5)** |
| --- | --- | --- | --- | --- | --- |
| **Pain and/or tenderness** |  |  |  |  |  |
| None | 32 (84.2%) | 9 (90.0%) | 4 (80.0%) | 14 (77.8%) | 5 (100.0%) |
| Grade 1: Mild | 5 (13.2%) | 1 (10.0%) | 1 (20.0%) | 3 (16.7%) | . |
| Grade 2: Moderate | 1 (2.6%) | . | . | 1 (5.6%) | . |
| **Erythema/redness** |  |  |  |  |  |
| None | 36 (94.7%) | 10 (100.0%) | 3 (60.0%) | 18 (100.0%) | 5 (100.0%) |
| Grade 1: Mild | 2 (5.3%) | . | 2 (40.0%) | . | . |
| **Induration/swelling** |  |  |  |  |  |
| None | 36 (94.7%) | 10 (100.0%) | 3 (60.0%) | 18 (100.0%) | 5 (100.0%) |
| Grade 1: Mild | 2 (5.3%) | . | 2 (40.0%) | . | . |
| **Erythema and/or Induration** |  |  |  |  |  |
| None | 34 (89.5%) | 10 (100.0%) | 1 (20.0%) | 18 (100.0%) | 5 (100.0%) |
| Grade 1: Mild | 4 (10.5%) | . | 4 (80.0%) | . | . |
| **Fever** |  |  |  |  |  |
| None | 32 (84.2%) | 8 (80.0%) | 5 (100.0%) | 17 (94.4%) | 2 (40.0%) |
| Grade 1: Mild | 6 (15.8%) | 2 (20.0%) | . | 1 (5.6%) | 3 (60.0%) |
| **Sleepiness/lethargy** |  |  |  |  |  |
| None | 29 (76.3%) | 7 (70.0%) | 4 (80.0%) | 15 (83.3%) | 3 (60.0%) |
| Grade 1: Mild | 8 (21.1%) | 3 (30.0%) | 1 (20.0%) | 3 (16.7%) | 1 (20.0%) |
| Grade 2: Moderate | 1 (2.6%) | . | . | . | 1 (20.0%) |
| **Rash** |  |  |  |  |  |
| None | 28 (73.7%) | 8 (80.0%) | 3 (60.0%) | 12 (66.7%) | 5 (100.0%) |
| Grade 1: Mild | 10 (26.3%) | 2 (20.0%) | 2 (40.0%) | 6 (33.3%) | . |
| **Vomiting** |  |  |  |  |  |
| None | 34 (89.5%) | 10 (100.0%) | 4 (80.0%) | 17 (94.4%) | 3 (60.0%) |
| Grade 1: Mild | 4 (10.5%) | . | 1 (20.0%) | 1 (5.6%) | 2 (40.0%) |
| **Anorexia** |  |  |  |  |  |
| None | 32 (84.2%) | 9 (90.0%) | 5 (100.0%) | 13 (72.2%) | 5 (100.0%) |
| Grade 1: Mild | 6 (15.8%) | 1 (10.0%) | . | 5 (27.8%) | . |
| **Seizure** |  |  |  |  |  |
| None | 38 (100.0%) | 10 (100.0%) | 5 (100.0%) | 18 (100.0%) | 5 (100.0%) |

For a given sign or symptom, each participant’s reactogenicity is counted once under the maximum severity across all study product administration visits.

Pain/tenderness, erythema, and induration symptoms are considered local reactogenicity if onset was within 7 full days following study product administration.

For erythema and induration, at each assessment time, the study site records the maximum perpendicular dimensions at the injection site.

Erythema and/or Induration is the maximum of the individual variables Erythema and Induration.

Symptoms are considered systemic reactogenicity if onset was within 7 full days following study product administration.

**Supplemental Table 5. Summary of adverse events, regardless of relationship to study product.** Number and percent of enrolled participants with each event of a given grade, overall and by arm.

|  |  | **Placebo (N=10)** | | | **20ug CH505TF gp120**  **2.5ug GLA-SE**  **(N=5)** | | | **20ug CH505TF gp120**  **5ug GLA-SE**  **(N=18)** | | | **5ug CH505TF gp120**  **5ug GLA-SE**  **(N=5)** | | |
| --- | --- | --- | --- | --- | --- | --- | --- | --- | --- | --- | --- | --- | --- |
| **Event**  **n (%)** | **Overall (N=38)** | **Grade 1** | **Grade 2** | **Grade 3** | **Grade 1** | **Grade 2** | **Grade 3** | **Grade 1** | **Grade 2** | **Grade 3** | **Grade 1** | **Grade 2** | **Grade 3** |
| **Any AE** | 38 (100%) | 1 (10.0%) | 5 (50.0%) | 4 (40.0%) | 2 (40.0%) | 2 (40.0%) | 1 (20.0%) | 5 (31.2%) | 8 (44.4%) | 5 (27.8%) | 2 (40.0%) | 3 (60.0%) | . |
| **Blood and lymphatic system disorders** |  |  |  |  |  |  |  |  |  |  |  |  |  |
| Lymphadenitis | 1 (2.6%) | . | . | 1 (10.0%) | . | . | . | . | . | . | . | . | . |
| **Congenital, familial and genetic disorders** |  |  |  |  |  |  |  |  |  |  |  |  |  |
| Dermoid cyst | 1 (2.6%) | . | . | . | . | . | . | . | . | . | . | 1 (20.0%) | . |
| **Eye disorders** |  |  |  |  |  |  |  |  |  |  |  |  |  |
| Eye discharge | 1 (2.6%) | . | . | . | . | . | . | . | . | . | . | 1 (20.0%) | . |
| **Gastrointestinal disorders** |  |  |  |  |  |  |  |  |  |  |  |  |  |
| Constipation | 1 (2.6%) | . | . | . | . | . | . | 1 (5.6%) | . | . | . | . | . |
| Diarrhoea | 12 (31.6%) | 7 (70.0%) | . | . | 2 (40.0%) | . | . | 2 (11.1%) | . | . | 1 (20.0%) | . | . |
| Mouth ulceration | 1 (2.6%) | . | . | . | . | . | . | . | 1 (5.6%) | . | . | . | . |
| Post-tussive vomiting | 1 (2.6%) | . | . | . | . | . | . | 1 (5.6%) | . | . | . | . | . |
| Stomatitis | 1 (2.6%) | . | 1 (10.0%) | . | . | . | . | . | . | . | . | . | . |
| Teething | 3 (7.9%) | 1 (10.0%) | . | . | 2 (40.0%) | . | . | . | . | . | . | . | . |
| Umbilical hernia | 1 (2.6%) | . | . | . |  | . | . | . | . | . | 1 (20.0%) | . | . |
| Vomiting | 4 (10.5%) | 3 (30.0%) | . | . | 1 (20.0%) | . | . | . | . | . | . | . | . |
| **General disorders and administration site conditions** |  |  |  |  |  |  |  |  |  |  |  |  |  |
| Hypothermia | 1 (2.6%) | . | . | . | . | . | . | 1 (5.6%) | . | . | . | . | . |
| Inflammation | 1 (2.6%) | . | . | . | . | . | . | . | . | . | 1 (20.0%) | . | . |
| Influenza like illness | 3 (7.9%) | . | 1 (10.0%) | . | . | . | . | . | 1 (5.6%) | . | . | 1 (20.0%) | . |
| Injection site swelling | 1 (2.6%) | . | 1 (10.0%) | . | . | . | . | . | . | . | . | . | . |
| Pyrexia | 1 (2.6%) | . | 1 (10.0%) | . | . | . | . | . | . | . | . | . | . |
| **Immune system disorders** |  |  |  |  |  |  |  |  |  |  |  |  |  |
| Hypersensitivity | 1 (2.6%) | . | . | . | 1 (20.0%) | . | . | . | . | . | . | . | . |
| Seasonal allergy | 1 (2.6%) | . | . | . | . | . | . | 1 (5.6%) | . | . | . | . | . |
| **Infections and infestations** |  |  |  |  |  |  |  |  |  |  |  |  |  |
| Hypersensitivity | 1 (2.6%) | . | . | . | 1 (10.0%) | . | . | . | . | . | . | . | . |
| Seasonal allergy | 1 (2.6%) | . | . | . | . | . | . | . | . | . | . | . | . |
| Body tinea | 3 (7.9%) | 1 (10.0%) | . | . | 1 (20.0%) | . | . | . | . | . | 1 (20.0%) | . |  |
| Bronchiolitis | 1 (2.6%) | . | . | . | . | . | . | . | . | 1 (5.6%) | . | . |  |
| Bronchitis | 1 (2.6%) | . | . | . | . | . | . | . | 1 (5.6%) | . | . | . | . |
| Conjunctivitis | 3 (7.9%) | 1 (10.0%) | 1 (10.0%) | . | . | . | . | 1 (5.6%) | . | . | . | . | . |
| Cytomegalovirus hepatitis | 1 (2.6%) | 1 (10.0%) | . | . | . | . | . | . | . | . | . | . | . |
| Ear infection | 1 (2.6%) | . | . | . | . | . | . | . | 1 (5.6%) | . | . | . | . |
| Fungal skin infection | 2 (5.3%) | . | . | 1 (10.0%) | . | . | . | . | 1 (5.6%) | . | . | . | . |
| Gastroenteritis | 9 (23.7%) | 2 (20.0%) | 1 (10.0%) | . | . | 1 (20.0%) | . | 2 (11.1%) | 2 (11.1%) | . | . | 1 (20.0%) | . |
| Gingivitis | 1 (2.6%) | . | . | . | . | . | . | . | 1 (5.6%) | . | . | . | . |
| Impetigo | 2 (5.3%) | . | 1 (10.0%) | . | . | . | . | 1 (5.6%) | . | . | . | . | . |
| Injection site abscess | 1 (2.6%) | 1 (10.0%) | . | . | . | . | . |  | . | . | . | . | . |
| Intestinal sepsis | 1 (2.6%) | . | . | . | . | . | . |  | . | 1 (5.6%) | . | . | . |
| Lower respiratory tract infection | 10 (26.3%) | 1 (10.0%) | 2 (20.0%) | . | 1 (20.0%) | . | . | 2 (11.1%) | 1 (5.6%) | 1 (5.6%) | . | 2 (40.0%) | . |
| Nasopharyngitis | 4 (10.5%) | 1 (10.0%) | . | . | 1 (20.0%) | . | . | 1 (5.6%) | . | . | 1 (20.0%) | . | . |
| Ophthalmia neonatorum | 1 (2.6%) | . | . | . | . | . | . | . | . | 1 (5.6%) | . | . | . |
| Otitis externa | 1 (2.6%) | . | . | . | . | . | . | . | 1 (5.6%) | . | . | . | . |
| Otitis media | 2 (5.3%) | . | . | . | . | . | . | . | 1 (5.6%) | . | . | 1 (20.0%) | . |
| Pharyngitis | 1 (2.6%) | . | 1 (10.0%) |  | . | . | . | . | . | . | . | . | . |
| Pneumonia | 2 (5.3%) | . | . | 1 (10.0%) | . | . | . | . | 1 (5.6%) | . | . | . | . |
| Tonsillitis | 4 (10.5%) | . | . | . | 1 (20.0%) | . | . | . | . | . | . | 3 (60.0%) | . |
| Upper respiratory tract infection | 18 (47.4%) | 1 (10.0%) | 5 (50.0%) | . | 2 (40.0%) | 2 (40.0%) | . | . | 5 (27.8%) | . | 3 (60.0%) | . | . |
| **Injury, poisoning and procedural complications** |  |  |  |  |  |  |  |  |  |  |  |  |  |
| Arthropod bite | 1 (2.6%) | 1 (10.0%) | . | . | . | . | . | . | . | . | . | . | . |
| Burns second degree | 1 (2.6%) | . | . | . | . | . | . | . | . | . | 1 (20.0%) | . | . |
| Thermal burn | 2 (5.3%) | 1 (10.0%) | 1 (10.0%) | . | . | . | . | . | . | . | . | . | . |
| **Investigations** |  |  |  |  |  |  |  |  |  |  |  |  |  |
| Alanine aminotransferase increased | 2 (5.3%) | 1 (10.0%) | . | 1 (10.0%) | . | . | . | . | . | . | . | . | . |
| Aspartate aminotransferase increased | 1 (2.6%) | . | 1 (10.0%) | . | . | . | . | . | . | . | . | . | . |
| Blood creatinine increased | 3 (7.9%) | 1 (10.0%) | . | . | . | . | . | 1 (5.6%) | . | . | 1 (20.0%) | . | . |
| Haemoglobin decreased | 30 (78.9%) | 4 (40.0%) | 5 (50.0%) | . | 2 (40.0%) | 1 (20.0%) | . | 12 (66.7%) | 2 (11.1%) | . | 3 (60.0%) | 1 (20.0%) | . |
| Neutrophil count decreased | 13 (34.2%) | 4 (40.0%) | 1 (10.0%) | 1 (10.0%) | 1 (20.0%) | . | 1 (20.0%) | 1 (5.6%) | 2 (11.1%) | 2 (11.1%) | . | . | . |
| Platelet count decreased | 1 (2.6%) | . | . | . | . | . | . | 1 (5.6%) | . | . | . | . | . |
| **Metabolism and nutrition disorders** |  |  |  |  |  |  |  |  |  |  |  |  |  |
| Decreased appetite | 3 (7.9%) | 1 (10.0%) | . | . | 1 (20.0%) | . | . | . | . | . | 1 (20.0%) | . | . |
| Hyperkalaemia | 1 (2.6%) | . | . | . | . | . | . | . | 1 (5.6%) | . | . | . | . |
| Underweight | 9 (23.7%)) | 4 (40.0%) | . | . | 2 (40.0%) | . | . | 3 (16.7%) | . | . | . | . | . |
| **Pregnancy, puerperium and perinatal conditions** |  |  |  |  |  |  |  |  |  |  |  |  |  |
| Jaundice neonatal | 1 (2.6%) | 1 (10.0%) | . | . | . | . | . | . | . | . | . | . | . |
| **Respiratory, thoracic and mediastinal disorders** |  |  |  |  |  |  |  |  |  |  |  |  |  |
| Bronchospasm | 1 (2.6%) | . | . | . | 1 (20.0%) | . | . | . | . | . | . | . | . |
| Cough | 8 (21.1%) | 3 (30.0%) | . | . | 2 (40.0%) | . | . | 2 (11.1%) | . | . | . | 1 (20.0%) | . |
| Nasal congestion | 9 (23.7%) | 3 (30.0%) | . | . | 1 (20.0%) | . | . | 4 (22.2%)) | . | . | 1 (20.0%) | . | . |
| Rhinorrhoea | 2 (5.3%) | 1 (10.0%) | . | . | 1 (20.0%) | . | . | . | . | . | . | . | . |
| **Skin and subcutaneous tissue disorders** |  |  |  |  |  |  |  |  |  |  |  |  |  |
| Dermatitis | 1 (2.6%) | 1 (10.0%) | . | . | . | . | . | . | . | . | . | . | . |
| Dermatitis allergic | 1 (2.6%) | . | . | . | . | . | . | 1 (5.6%) | . | . | . | . | . |
| Dermatitis contact | 1 (2.6%) | 1 (10.0%) | . | . | . | . | . | . | . | . | . | . | . |
| Dermatitis diaper | 8 (21.1%) | 2 (20.0%) | . | . | 1 (20.0%) | . | . | 4 (22.2%) | 1 (5.6%) | . | . | . | . |
| Dry skin | 1 (2.6%) | . | . | . | . | . | . | 1 (5.6%) | . | . | . | . | . |
| Eczema | 4 (10.5%) | 2 (20.0%) | . | . | 1 (20.0%) | . | . |  | . | . | 1 (20.0%) | . | . |
| Intertrigo | 1 (2.6%) | . | . | . | 1 (20.0%) | . | . |  | . | . | . | . | . |
| Miliaria | 1 (2.6%) | . | . | . | . | . | . | . | . | . | 1 (20.0%) | . | . |
| Rash | 10 (26.3%) | 2 (20.0%) | 1 (10.0%) | . | 2 (40.0%) | . | . | 4 (22.2%) | . | . | 1 (20.0%) | . | . |
| Rash papular | 2 (5.3%) | 1 (10.0%) | . | . | . | . | . | 1 (5.6%) | . | . | . | . | . |
| Rash vesicular | 1 (2.6%) | . | . | . | . | . | . | . | 1 (5.6%) | . | . | . | . |
| Seborrhoeic dermatitis | 3 (7.9%) | 2 (20.0%) | . | . | . | . | . | . | 1 (5.6%) | . | . | . | . |
| Skin induration | 1 (2.6%) | . | . | . | . | . | . | 1 (5.6%) | . | . | . | . | . |

N is the number of participants who experienced one or more AEs of a specific severity within a body system or MedDRA preferred term. Percentages are the number of participants who experienced an event of a specific severity divided by the number enrolled. For participants with multiple events within a body system or for a MedDRA preferred term, the maximum severity is counted.

**Supplemental Table 6. Summary of adverse events deemed related to study product(s).** Number and percent of enrolled participants who experienced a given related event, overall and by arm.

|  | | **Placebo (N=10)** | | | | **CH505TF/GLA-SE vaccine (N=28)** | | | |
| --- | --- | --- | --- | --- | --- | --- | --- | --- | --- |
| **Event**  **N (%)** | **Overall**  **(N=38)** | **Grade 1** | **Grade 2** | **Grade 3** | **Grade 4** | **Grade 1** | **Grade 2** | **Grade 3** | **Grade 4** |
| Neutrophil count decreased | 1 (2.6%) | 1 (10.0%) | 0 (0.0%) | 0 (0.0%) | 0 (0.0%) | 0 (0.0%) | 0 (0.0%) | 0 (0.0%) | 0 (0.0%) |

N is the number of participants who experienced one or more AEs of a specific relationship to study product within a body system or MedDRA preferred term. Percentages are the number of participants with an event of a specific relationship divided by the number enrolled. For participants with multiple events within a body system or for a MedDRA preferred term, the maximum severity is counted.

**­**
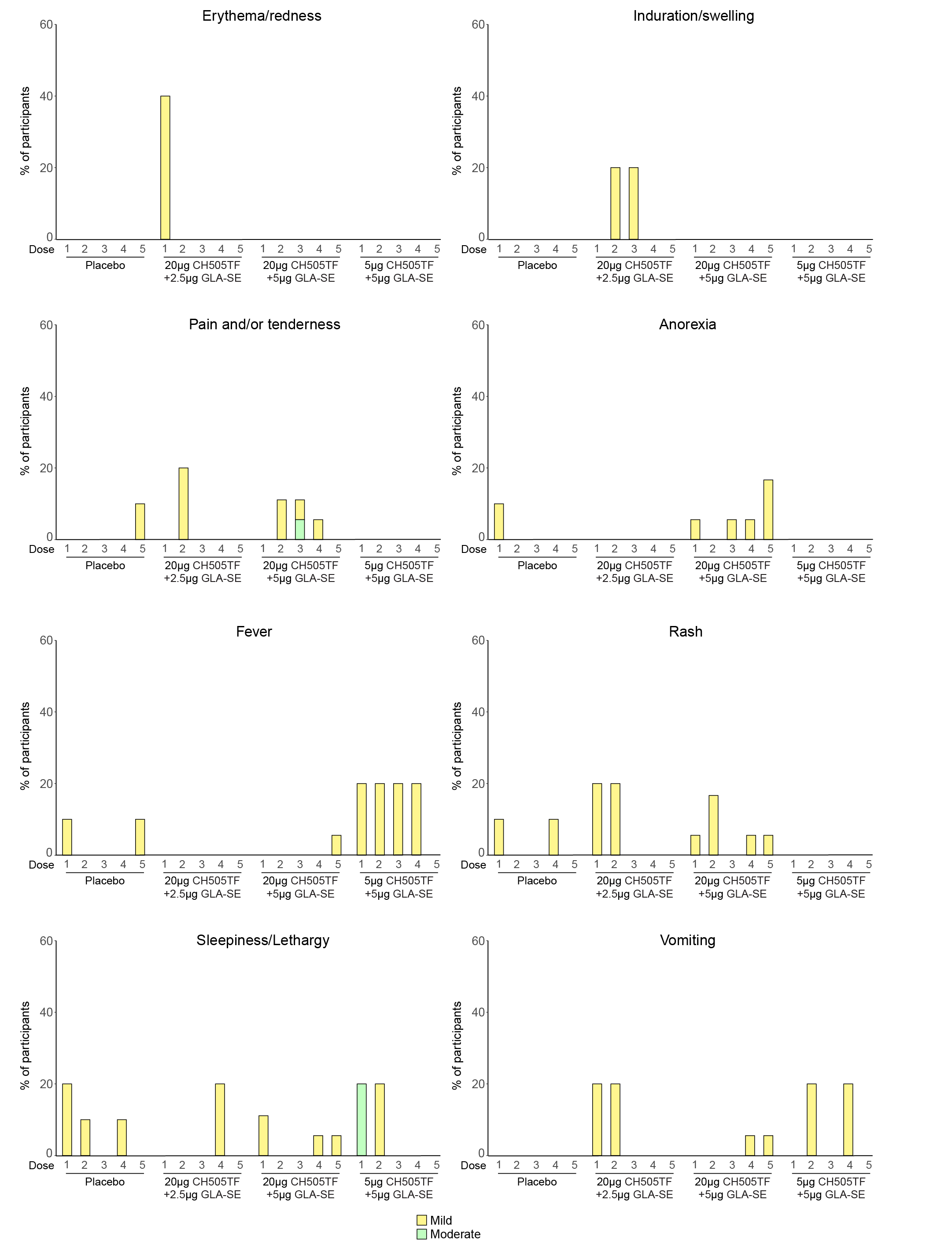

**Supplemental Figure 1. Systemic and local reactogenicity.** Percent of enrolled infants experiencing each type of reactogenicity, by arm, injection number and severity.

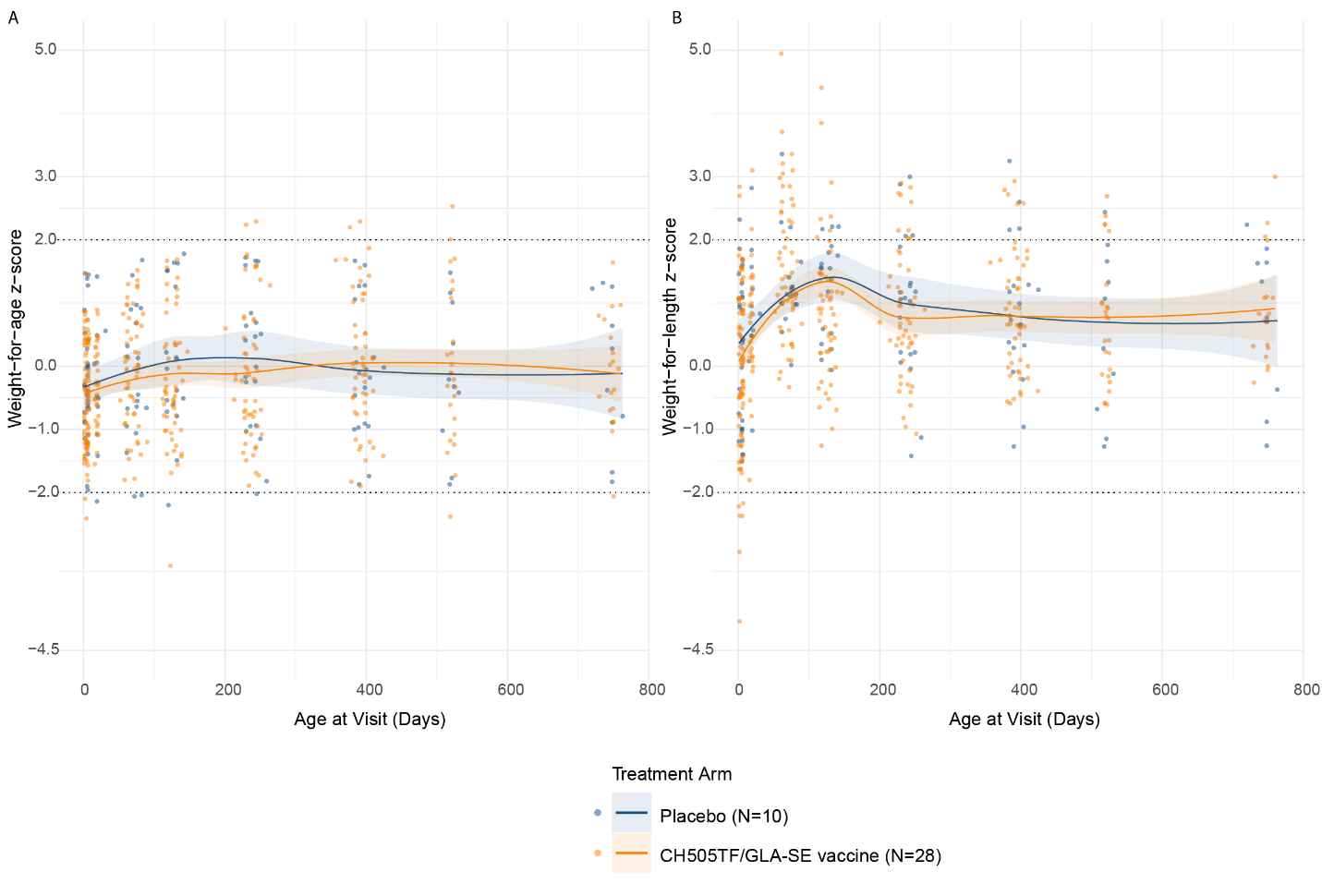

**Supplemental Figure 2. Anthropometric measures over follow-up.** Weight-for-Age z-score (WAZ) and Weight-for-Length z-score (WLZ) for all enrolled participants by arm. Each line indicates the sample mean. The shading indicates a pointwise 95% CI. WAZ is a measure of a child's weight in relation to their age, based on the distribution of weight in a reference population (*1*). The z-score is defined as: (Actual Weight (W)−Median Weight for Age (M))/ Standard Deviation (SD), where Standard Deviation (SD) represents the variability or spread of weights in the reference population. WLZ measures a child's weight in relation to their length or height, based on the distribution of weight and length in a reference population. The WLZ z-score is defined as: (Actual Weight (W)−Median Weight for Length (M))/ Standard Deviation (SD), where Standard Deviation (SD) represents the variability or spread of weight in a reference population.

1. WHO, Training course on child growth assessment (2008). <https://iris.who.int/bitstream/handle/10665/43601/9789241595070_A_eng.pdf?sequence=1&isAllowed=y>.
